# Linking Cognitive Variability and Alzheimers Disease Biomarkers by Neurocognitive Status

**DOI:** 10.64898/2026.01.30.26345235

**Authors:** Shayne S.-H. Lin, Alicia Milam, Andrew M. Kiselica, Stephen L. Aita, Mubarick Saeed, Troy Webber, Steven Paul Woods, Nicholas C. Borgogna, Keenan A. Walker, Vidyulata Kamath, Kristina Visscher, Charles F. Murchison, David S. Geldmacher, Erik D. Roberson, Benjamin D. Hill, Victor A. Del Bene

## Abstract

**Objective:** To assess intra-individual cognitive variability (IICV) in relation to Alzheimer’s Disease (AD) biomarkers.

**Methods:** The sample included 879 adults from the National Alzheimer’s Coordinating Center, aged 50 and above with a complete neuropsychological evaluation and AD biomarker data available (64% cognitively intact; 36% cognitively impaired). We conducted a series of moderated regression models where AD biomarkers, neurocognitive status, and their interaction effects predicted IICV. IICV measures included demographically adjusted normed scores for the intraindividual standard deviation (iSD) and coefficient of variance (CoV). AD biomarkers included cerebrospinal fluid (CSF) measures of Aβ_1-42_, phosphorylated tau 181 (p-Tau_181_), and total tau (t-Tau), as well as amyloid positron emission tomography (PET; with both continuous centiloid values and a dichotomous variable).

**Results:** Increased AD biomarker burden was associated with increased IICV among cognitively impaired individuals (correlational strength ranging from .206 to .391 for iSD and from .149 to .460 for CoV) but not among the cognitively intact group (correlational strength ranging from .008 to .085 for iSD and from .016 to .085 for CoV). The pattern of results held even after controlling for demographic factors and was comparable in magnitude to the association between AD biomarkers and mean cognitive performance.

**Conclusions:** Increases in measures of amyloid, soluble tau, and neurodegeneration are associated with increased IICV among cognitively impaired older adults. The findings underscore the potential of IICV as a sensitive outcome measure in the AD clinical disease phase. Future studies should replicate findings longitudinally and in more diverse samples.

## Introduction

Intra-individual cognitive variability (IICV) has emerged as a sensitive marker of cognitive decline associated with Alzheimer’s Disease (AD) pathology^1,2^. IICV quantifies the variation in cognitive performance within an individual, either across multiple trials within a test (referred to as Consistency) or across different tests or scores within a neuropsychological test battery (referred to as Dispersion) in a single testing session^3,4^. For the current study, we use the term IICV to refer to Dispersion of scores.

IICV differentiates normal cognitive aging from pathological aging due to Alzheimer’s Disease AD^1^. From a diagnostic perspective, relative to healthy controls, older adults with mild cognitive impairment (MCI) and dementia due to AD yield a neuropsychological profile characterized by increased IICV^2^. Longitudinally, heightened IICV at baseline predicts the transition from normal aging to MCI and from MCI to AD^5,6^. IICV is also associated with cognitive and functional impairment cross-sectionally and predicts cognitive and functional decline longitudinally^7–9^. Finally, IICV is linked to several biomarkers of AD , including grey matter atrophy and cortical thinning of signature AD-related brain regions (e.g., entorhinal cortex, hippocampus, and the parahippocampal gyrus)^10^, functional disruption of the default mode and fronto-parietal executive control networks^11^, anatomical disruption of white matter integrity^12^, presence of the apolipoprotein E ε4 allele (a genetic risk factor for AD)^1,13^, as well as amyloid-beta (Aβ)^14^ and tau accumulation^15,16^. Thus, cumulative but preliminary evidence advances IICV as a sensitive outcome marker of AD-related diagnosis, cognitive and adaptive functioning, and neurobiological changes^6,9,15,16^.

Despite an expansion of the literature on IICV over the past five years, scientific understanding of IICV and its clinical application has yet to reach maturity. One major outstanding gap in the literature regarding IICV concerns whether there are unique windows of clinical disease severity in which IICV most closely tracks with AD neuropathology; that is, research has yet to clarify the phase of clinical progression in which IICV can be best used as a clinical outcome measure (e.g., in the preclinical versus clinical stage of disease). Addressing these gaps is a necessary step towards implementation of IICV as a clinical outcome measure. Therefore, this study comprehensively assessed the relationship of AD biomarkers (CSF, PET) and IICV as a function of the presence versus absence of cognitive impairment, expanding upon prior studies on the neuropathological correlates of IICV^14–16^. We assessed several AD biomarkers, including cerebrospinal fluid (CSF) measures of Aβ_1-42_, phosphorylated tau_181_ (p-Tau_181_), and total tau (t-Tau), as well as amyloid positron emission tomography (PET), in a large cohort. We hypothesized that demographically adjusted, normed IICV (with lower scores indicating higher variation) would be positively associated with CSF Aβ and negatively associated with PET Aβ, CSF p-Tau_181_, and CSF t-Tau levels. We further anticipated that this relationship would be moderated by neurocognitive status, such that the strongest effects would be observed among cognitively impaired individuals.

## Methods

### Sample

The third version of the Uniform Data Set (UDS 3.0) from the National Alzheimer’s Coordinating Center (NACC; https://naccdata.org/) was requested on March 26^th^, 2024. The NACC is housed at the University of Washington and collects data from 42 Alzheimer’s Disease Research Centers (ADRCs) from the United States. Detailed descriptions of the NACC and the UDS can be found elsewhere^17,18^. The current sample consisted of data from participants’ initial visit with data collection time ranging from the commencement of the UDS data collection in 2005 to the data freeze on March 2024.

See Figure 1 for a STROBE diagram depicting the sample selection process^19^. See Table 1 for demographic characteristics of the final sample (n = 879). Inclusion criteria were: 1) completion of the UDS 3.0 at baseline, 2) age 50 and beyond, 3) English as participants’ primary language and the language of administration, 4) the Global Clinical Dementia Rating (CDR) lower than 2^20^, 5) clear diagnostic impression of cognitive status (i.e., intact or presence of a neurocognitive disorder), 6) a presumed AD etiology for those with a neurocognitive disorder (i.e., MCI or dementia), 7) adequate hearing and vision for the purpose of testing, and 8) available data for key biomarker neuropsychological, and demographic variables. Of note, individuals with moderate-severe dementia were excluded due to the high likelihood of being unable to meaningfully participate in neuropsychological testing^20^.

**Figure 1.**
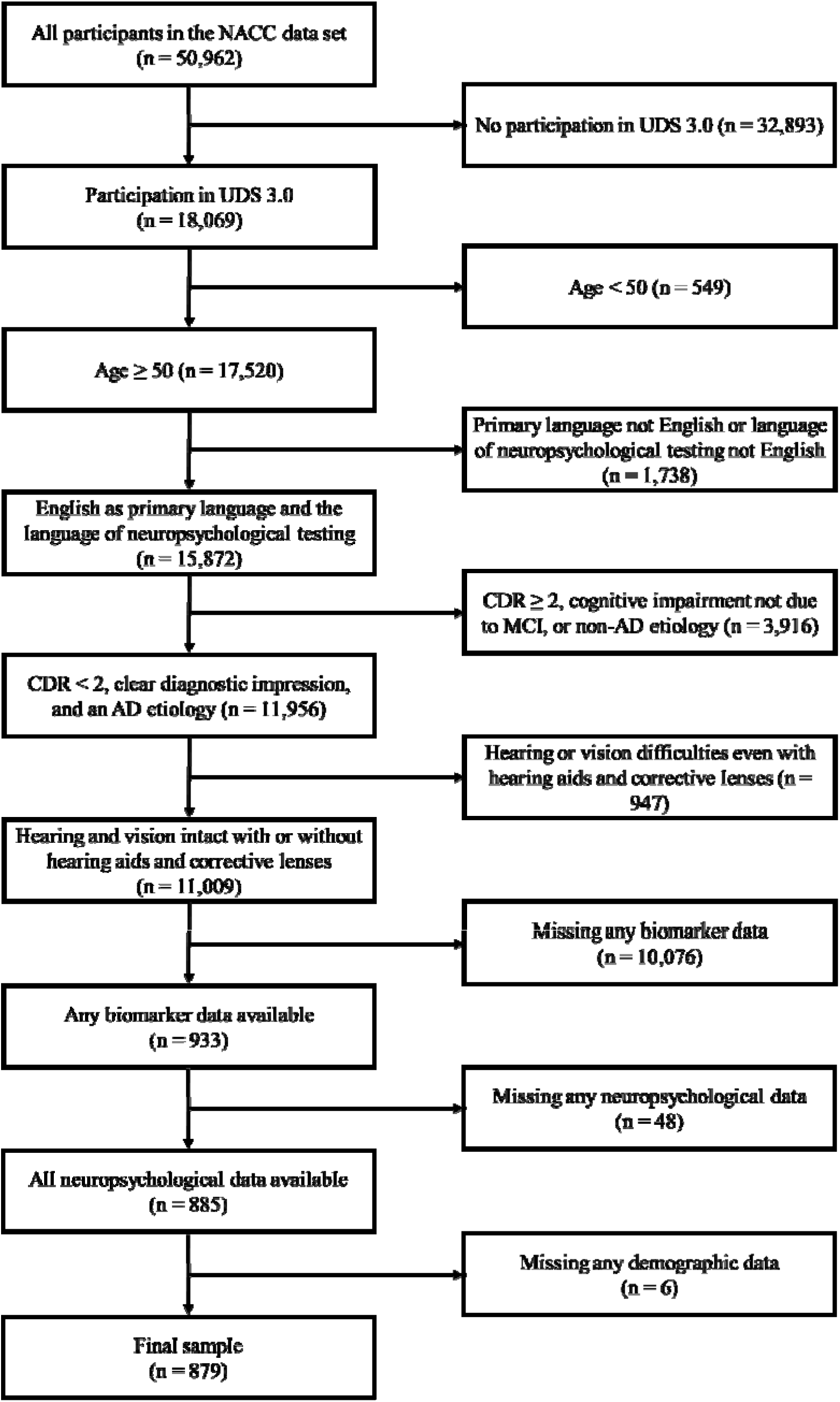
STROBE diagram describing sample selection process from the NACC data pool.

**Table 1.**
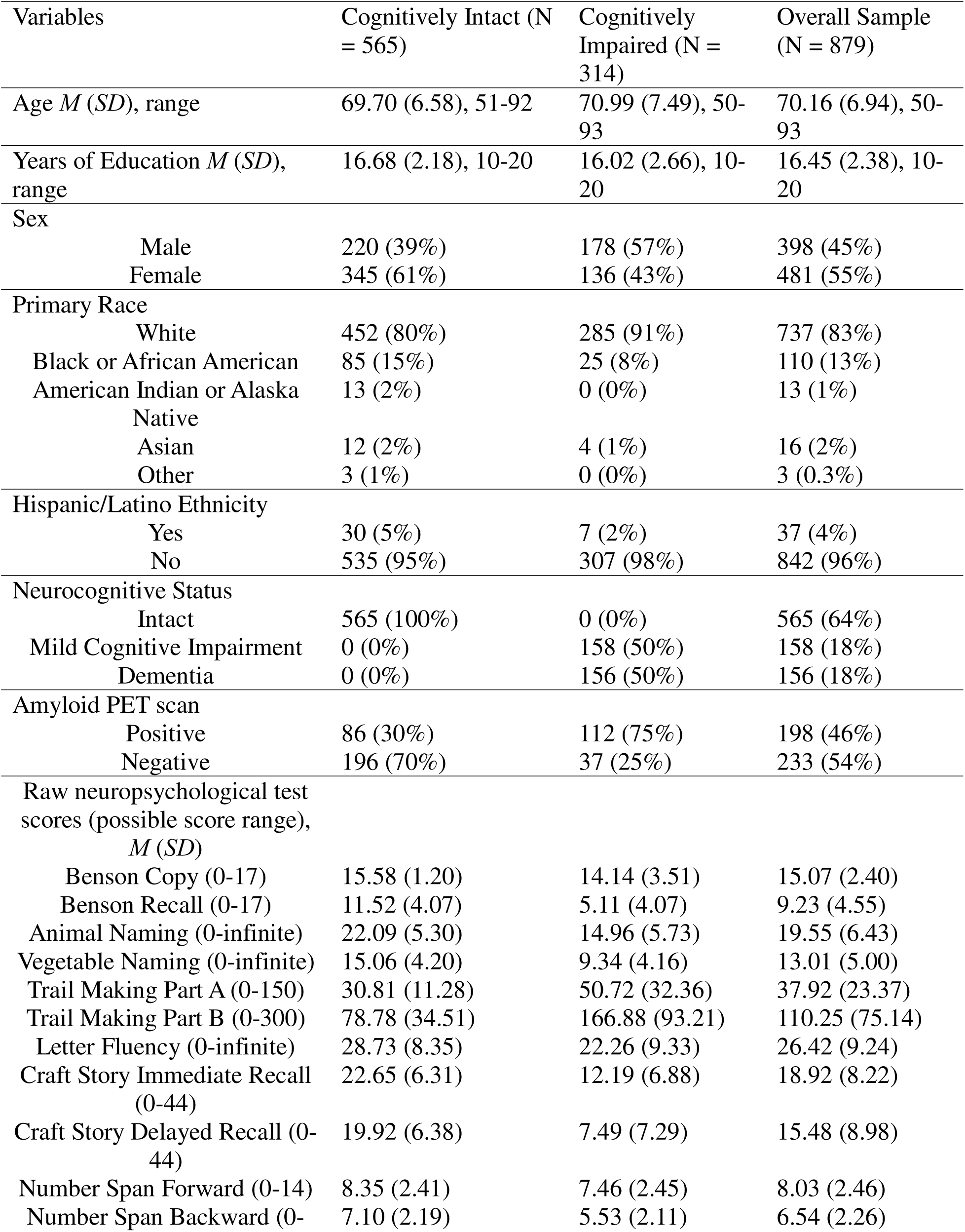
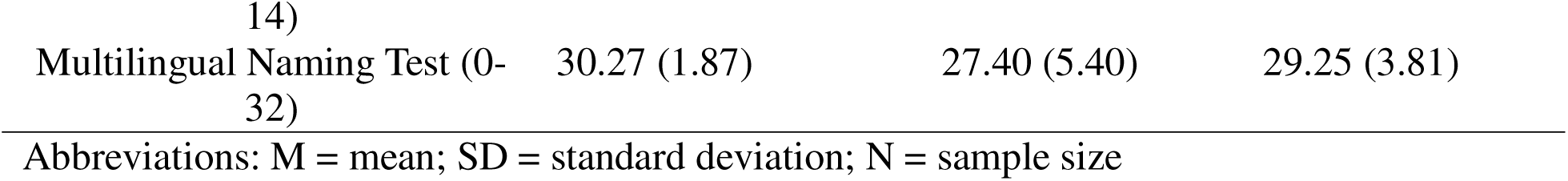
Sample Characteristics.

### Measures

#### IICV

The battery of neuropsychological tests from the UDS 3.0 included in the calculation of IICV are listed in Table 1^17,18,21^. For the individual tests, missing values due to verbal refusal, physical difficulties, and unknown reasons were removed from further analysis. Cases with missingness as a result of cognitive or behavioral difficulties (n = 40) were assigned with the lowest possible test scores, recognizing that cognitive impairment led to the discontinuation of test administration^22^. Specifically, the lowest scores for the Trial Making Test Parts A and B were set to 150 and 300 seconds, respectively. For all other tests, the lowest score was set to 0^21^. Raw neuropsychological test scores from each measure were transformed into demographically adjusted z-scores based on previously published norms^17,21,23^.

Two IICV scores, the intraindividual standard deviation (iSD) and coefficient of variance (CoV), were then calculated after transforming demographically adjusted z-scores into T-scores of for each individual test which avoid negative and zero values^17^. While iSD is conceptualized as the standard deviation among the T-scores of individual tests, CoV adjusts for global cognitive performance by dividing iSD by the mean T-score (i.e., overall test battery mean)^7,11,21^. Finally, iSD and CoV were then transformed to scaled scores before being converted into demographically adjusted z-scores, per recommendations from prior literature^21^. The demographically adjusted z-scores of IICV are interpreted such that higher scores indicate lower variability (i.e., lower z-scores indicate more variability).

### AD Biomarkers

AD biomarkers included Aβ_1-42_ (measures amyloid accumulation), t-Tau (measures neurodegeneration), and p-Tau_181_ (measures soluble tau accumulation) from CSF, as well as centiloid values and dichotomous Aβ status (positive versus negative) from PET scans (measures amyloid accumulation). For CSF, data were collected through lumbar puncture with the Enzyme-Linked Immunosorbent Assay Analytic Protocol or the Luminex Multi-Analyte Profiling. Assay technology was site-dependent, though all CSF data were assayed in one batch^24^. Values represent picogram per milliliter (pg/ml). Notably, given the positive skewness of t-Tau, this variable was initially log-transformed. However, as data remained in a non-normal distribution with skewness of -1.66 and Kurtosis of 4.27, square root transformation was attempted, resulting in distribution of t-Tau within normal limit, skewness = 0.41, Kurtosis = 1.36. Dichotomized variables were also established by a cutoff of ≤ 700 pg/ml for Aβ_1-42_, ≥ 60 pg/ml for p-Tau_181_, and ≥ 400 pg/ml for t-Tau^25^.

Amyloid PET data were retrieved from the Standardized Centralized Alzheimer’s & Related Dementias Neuroimaging dataset (SCAN; supported by the National Institute on Aging with the U24 grant [AG067418]), a project that standardized the acquisition, curation, and analysis of neuroimages obtained by multiple ADRCs. SCAN PET data were all defaced at the Aging and Dementia Imaging Research (ADIR) Laboratory at the Mayo Clinic with quality assessment provided by the University of Michigan. For a detailed description of the imaging processing protocol, visit https://scan.naccdata.org/. Two variables were derived from the amyloid PET scan: centiloid values and amyloid status. Briefly, centiloid is a standardized quantitative amyloid measure. Mathematically, centiloid is calculated by a linear transformation of the cortical summary of the standardized uptake value ratios (SUVRs) normalized to the whole size of the cerebellum. This process was completed by the Global Alzheimer’s Association Interactive Network at University of California, Berkeley.

A centiloid value of 0 represents the amyloid level of typical young adults, while 100 is anchored at the level of typical of patients with dementia due to AD, though values outside of the 0-100 bound are possible and common. Additionally, we analyzed a binary amyloid status value representing the presence versus absence of elevated amyloid in the brain. Given the inconsistency among ADRCs in their operationalization of this binary decision, two methods were employed to set the threshold: 1) converting an established MRI-dependent threshold to the SUVRs and 2) two standard deviations above the average SUVR of typical young adults^26^. For each type of PET scan radiotracer, both threshold methods were used in adjudicating the presence or absence of abnormally elevated cortical amyloid.

### Demographics and Diagnoses

Demographics of interest included age, sex, education, race, and ethnicity. Given the scarcity of participants of a minoritized backgrounds in this sample (see Table 1), individuals from racial and/or ethnical minority were aggregated into one group for analyses involving race (non-Hispanic White = 0, minorities = 1). Clinical diagnoses were dichotomized into a cognitively intact group and a cognitively impaired group, which consisted of individuals with consensus diagnoses of MCI or mild dementia.

### Data Analyses

We completed data analyses and visualizations using R version 4.4.2. The main goal for the current study was to examine the relation between AD biomarkers and IICV as moderated by neurocognitive status. To achieve this goal, regression analyses were conducted where AD biomarkers, including Aβ_1-42_, t-Tau, and p-tau_181_ from CSF, as well as centiloid and Aβ from amyloid-PET, were taken as independent variables to predict the two IICV scores (iSD and CoV) with neurocognitive status (intact = 0, impaired = 1) as a moderator. All AD biomarker indices were scaled for inferential statistical analyses. To avoid multicollinearity, all continuous AD biomarkers were standardized prior to statistical analysis and regressed onto each IICV index in separate models, and we reviewed results qualitatively to assess consistency of relations across AD biomarkers. Post-hoc correlation analyses were performed, stratified by neurocognitive status groups when there was a significant moderation effect (i.e., simple slopes analysis). Sensitivity analyses with demographics as covariates included in the regression models were explored to test the robustness of the statistical results. Additional sensitivity only the Black and African American subsample were conducted to test for the to this underrepresented group.

Exploratory analyses were also conducted with mean cognitive performance as a dependent variable to assess equivalency of results to those obtained using IICV . Finally, we examined IICV in the context of the categorical ATN ^25^ with dichotomized CSF Aβ (positive: A+; negative: A-), p-Tau_181_ (positive: T+; negative: T-), and t-Tau (positive: N+; negative: N-) as independent variables and IICV as a dependent variable in separate Analysis of Variance (ANOVA) models.

## Results

Bivariate correlations and descriptive statistics are reported in Table 2. All skewness and kurtosis were within normal limit. Both indices of IICV significantly correlated with all AD biomarkers, with most effect sizes in the moderate range (correlational strength ranging from *r* = 0.287 – 0.687). Group differences on key variables by neurocognitive status are shown in Table 3. The cognitively intact and the cognitively impaired groups differed on all key variables, including demographics, IICV, and AD biomarkers. The cognitively impaired group showed diminished demographically adjusted iSD and CoV (i.e., high IICV), reduced CSF amyloid-β, and increased CSF t-Tau and p-Tau_181_, amyloid PET centiloid values, and rates of amyloid PET positivity. In summary, associations consistently suggested that more AD neuropathology burden was associated with increased IICV.

**Table 2.**
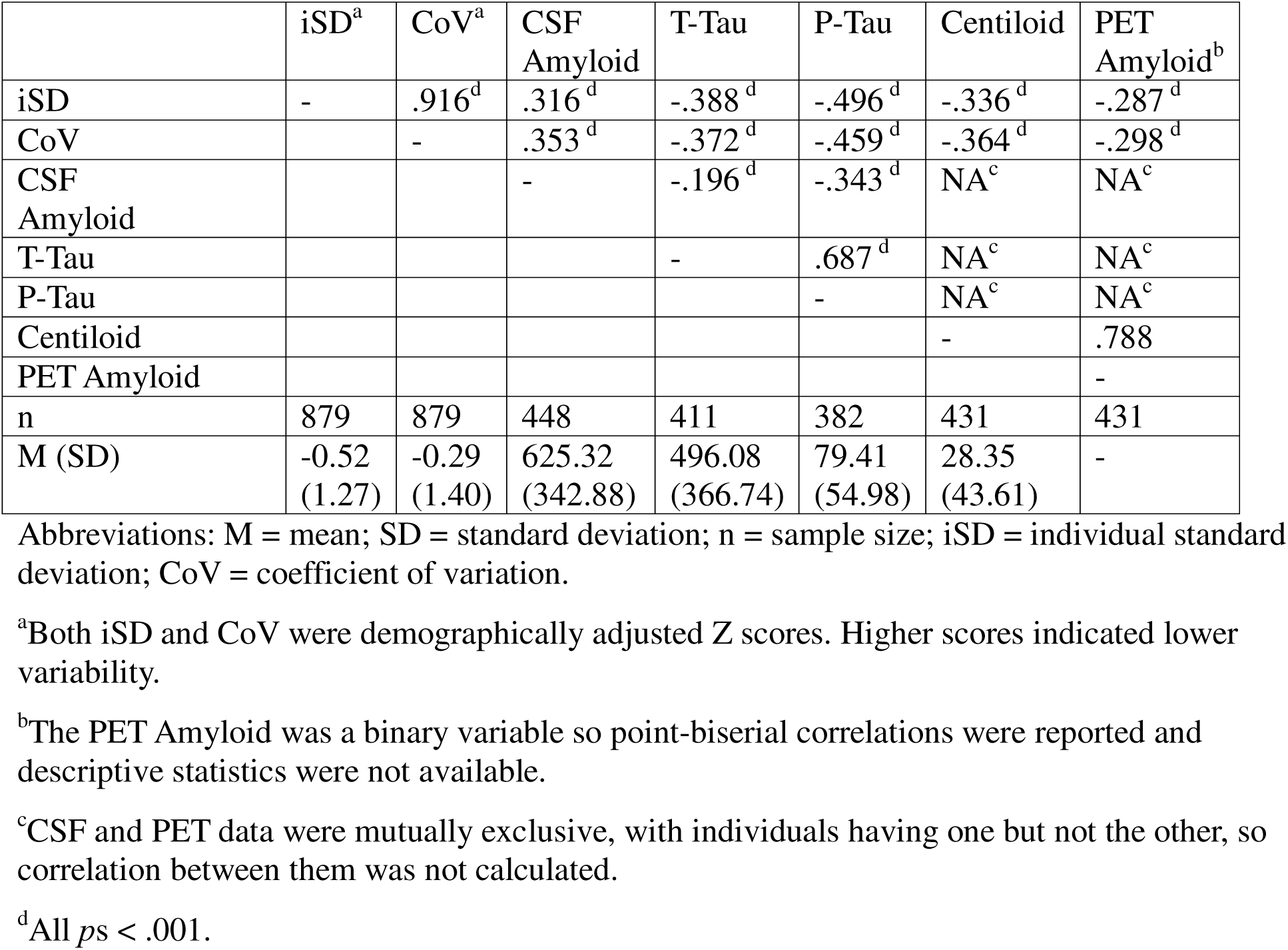
Correlations among Variables.

**Table 3.** Group Differences among Variables.

Group Differences among Variables
|  | Statistical Analyses<sup>a, c</sup> | Intact<sup>b</sup> | Impaired<sup>b</sup> | Effect Sizes<sup>a</sup> |
| --- | --- | --- | --- | --- |
| Age, M (SD) | -2.65** | 69.70 (6.58) | 70.99 (7.49) | 0.19 |
| Sex, n (%) | 24.95*** | M: 220 (39%)<br>F: 345 (61%) | M: 178 (57%)<br>F: 136 (43%) | 0.49 |
| Race, n (%) | 20.54*** | W: 427 (76%)<br>Non-W: 138 (24%) | W: 278 (89%)<br>Non-W: 36 (11%) | 0.40 |
| Ethnicity, n (%) | 4.02* | Non-H: 550 (95%)<br>H: 30 (5%) | Non-H: 307 (98%)<br>H: 7 (2%) | 0.03 |
| Education, M (SD) | 4.00*** | 16.68 (2.18) | 16.02 (2.66) | 0.28 |
| iSD<sup>d</sup>, M (SD) | 20.89*** | 0.02 (-0.04) | -1.50 (1.15) | 1.47 |
| CoV<sup>d</sup>, M (SD) | 24.82*** | 0.38 (0.97) | -1.50 (1.24) | 1.75 |
| CSF Amyloid, M (SD) | 9.48*** | 732.41 (344.22) | 441.63 (250.94) | 0.93 |
| T-Tau<sup>e</sup>, M (SD) | -11.07*** | 17.92 (6.82) | 25.6 (6.87) | 1.12 |
| P-Tau, M (SD) | -13.60*** | 54.28 (32.86) | 118.69 (59.49) | 1.43 |
| Centiloid, M (SD) | -12.30*** | 12.18 (30.95) | 58.95 (47.60) | 1.25 |
| PET Amyloid, M (SD) | 76.55*** | N: 196 (70%)<br>P: 86 (30%) | N: 37 (25%)<br>P: 112 (75%) | 6.85 |

Abbreviations: M = males; F = females; W = White; H = Hispanics; N = negative; P = positive.
<sup>a</sup>Chi-square and t tests were reported as statistical analyses and odds ratio and Cohen's d as effect sizes for categorical and continuous variables, respectively.
<sup>b</sup>For continuous variables, means and standard deviations (in parentheses) were reported while frequencies and percentages (in parentheses and anchored within columns) were reported for categorical variables.
<sup>c</sup>\* $p < .05$ , \*\*\* $p < .001$ .
<sup>d</sup>Both iSD and CoV are demographically adjusted Z scores. Higher scores indicate lower variability.
<sup>e</sup>t-Tau was square root transformed due to positive skewness.

In a regression model, we entered AD biomarkers, neurocognitive status, and their interaction term as independent variables in predicting iSD and CoV. These results are shown in Table 4. All variance inflation factors (VIFs) were within normal limits, and therefore, no multicollinearity was present. Across all AD biomarkers, the interactions with neurocognitive status were statistically significant in the prediction of iSD. Post-hoc correlation analyses confirmed the hypothesized moderation effect: the relationship between AD biomarkers and IICV was statistically significant in the cognitively impaired group but not for the cognitively intact group. For CoV, there was a similar pattern, though the moderation of neurocognitive status on CSF t-Tau and p-Tau_181_ and amyloid positivity on PET did not reach statistical significance. Post-hoc correlations and interaction visualizations stratified by neurocognitive status group can be found in Table 5 and Figure 2.

**Figure 2.**
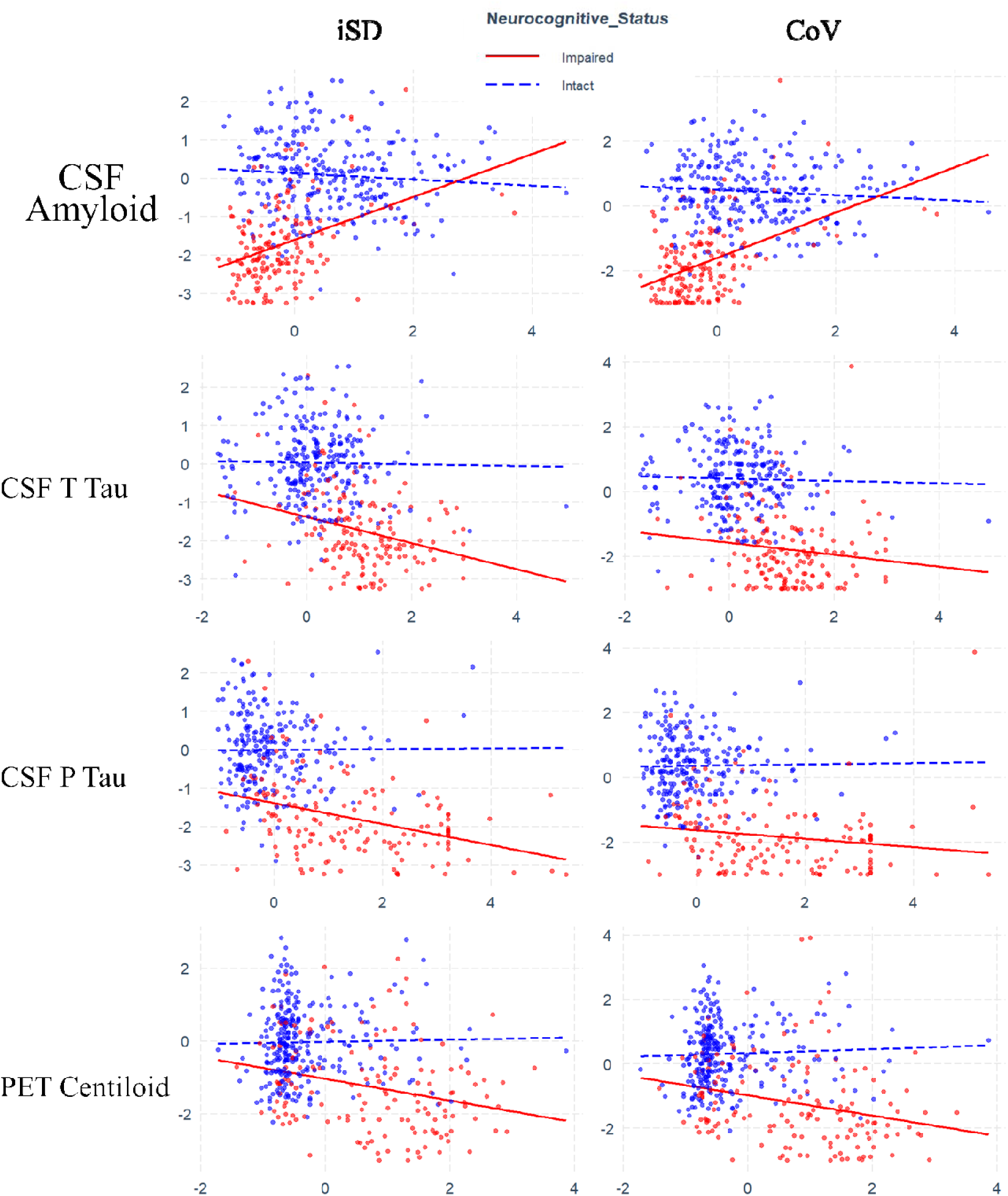
Interactions plots depicting the moderation between neurocognitive status and Alzheimer’s Disease biomarkers on Intra-Individual Cognitive Variability. X axis represents IICV (either iSD or CoV) while Y axis shows AD biomarkers. Both iSD and CoV are demographically adjusted Z scores. Higher scores indicate lower variability. AD biomarkers are also scaled.

**Figure 3.**
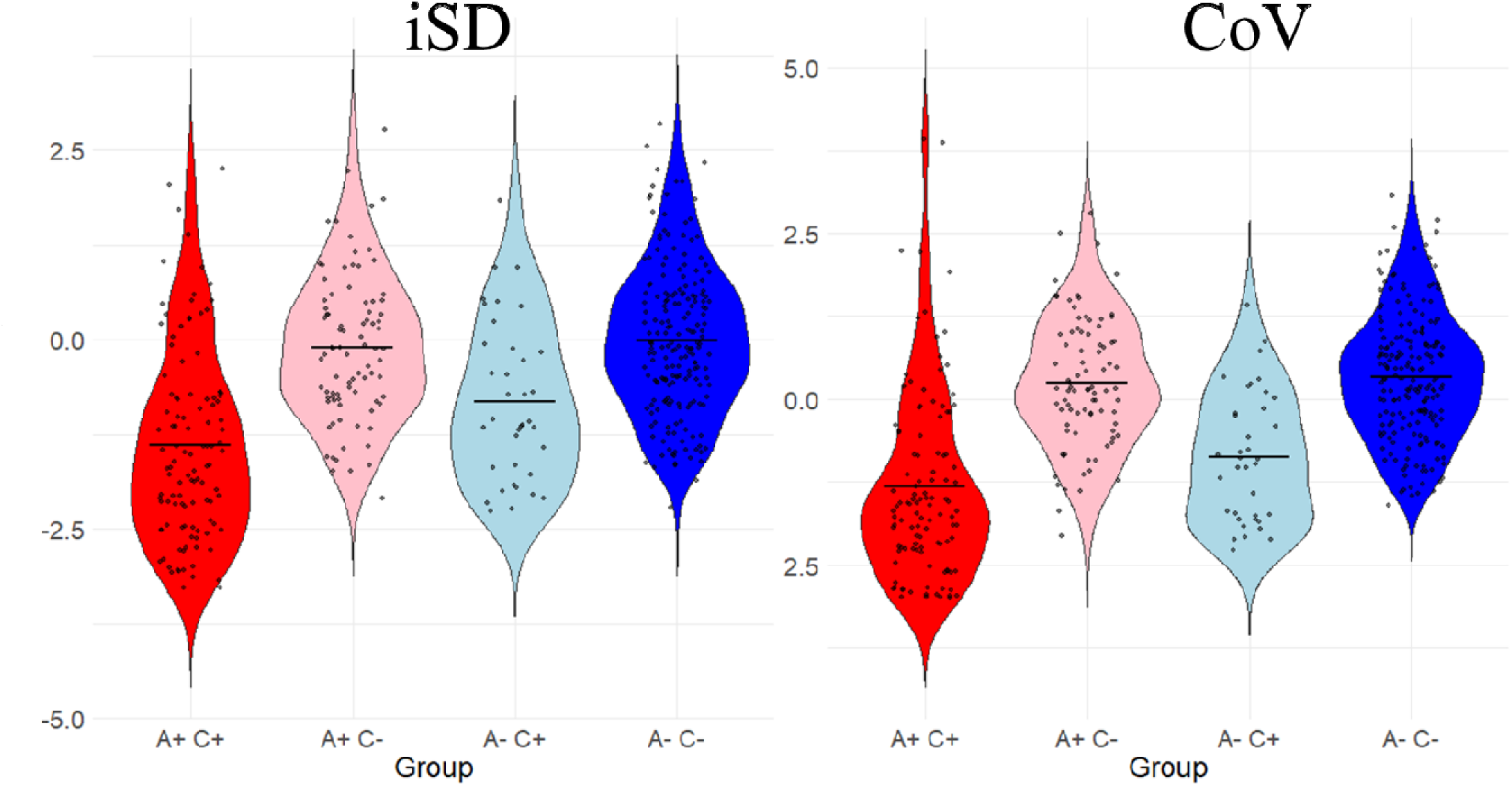
The interactions between neurocognitive status (C+ cognitively impaired; C-cognitively intact) and PET Amyloid status (A+ Amyloid positive; A- Amyloid negative) on demographically adjusted Z scores of individual standard deviation (iSD) and coefficient of variation (CoV). The horizontal straight lines represent the means for each group. Both iSD and CoV were demographically adjusted Z scores. Higher scores indicated lower variability.

**Table 4.**
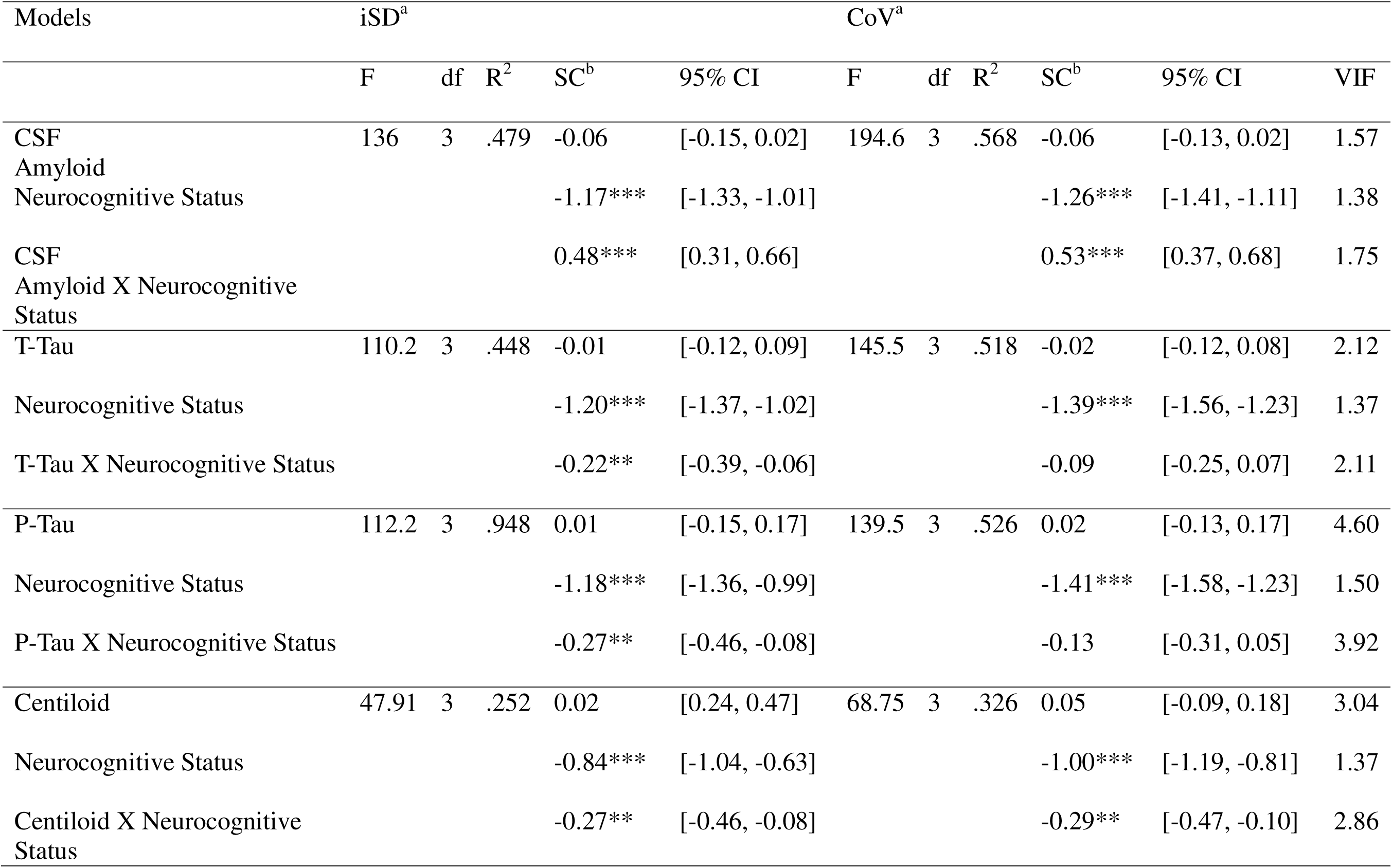
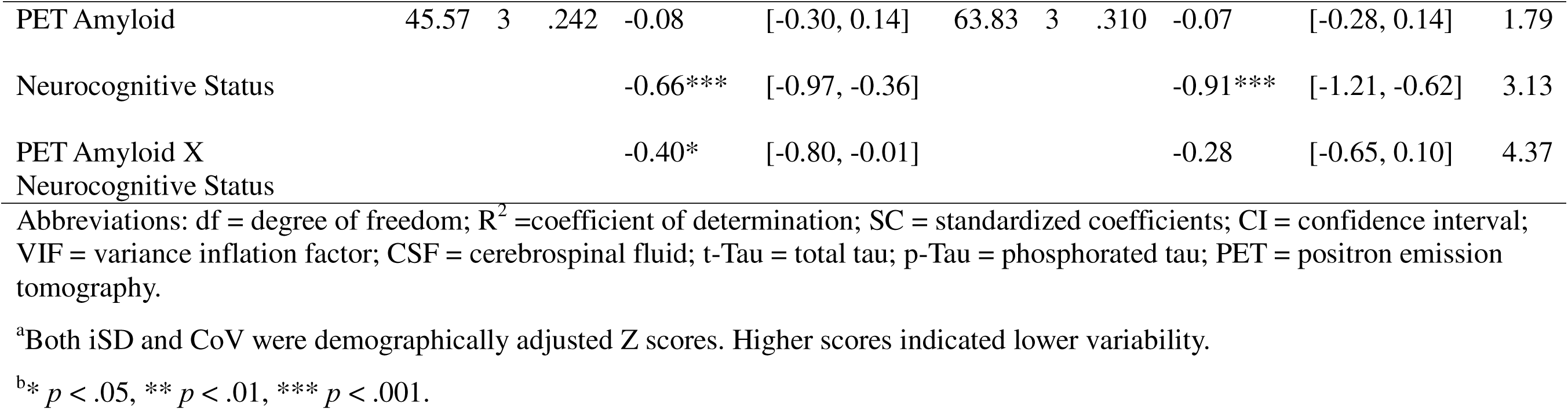
Interactions between Biomarkers and Neurocognitive Status in the Prediction of IIV.

**Table 5.** Post-hoc Correlation Analysis.

|  | ISD<sup>a</sup> |  | CoV<sup>a</sup> |  |
| --- | --- | --- | --- | --- |
|  | Intact | Impaired | Intact | Impaired |
| CSF Amyloid | -.085 | .391*** | -.085 | .460*** |
| T-Tau | -.017 | -.269*** | -.030 | -.136 |
| P-Tau | .008 | -.362*** | .016 | -.161* |
| Centiloid | .022 | -.266** | .045 | -.260** |
| PET Amyloid<sup>b</sup> | .045 | -.206* | -.043 | -.149 |

<sup>a</sup>Both iSD and CoV were demographically adjusted Z scores. Higher scores indicated lower variability.
<sup>b</sup>Point-biserial correlations were reported for PET Amyloid.

Additional sensitivity analyses were employed to examine the robustness of the study results. After accounting for demographic covariates, the primary findings were consistent. Notably, most demographic variables were not associated with the demographically adjusted z-scores, showing the effectiveness of demographic correction on normative cognitive test performance, as shown in Supplemental Material Table 1. Within Black and African American participants (*n* = 85 for cognitively intact and *n* = 25 for cognitively impaired), both IICV indices were only statistically significantly associated with p-Tau_181_ and no other AD biomarkers (Supplemental Material Table 2). None of the interaction terms between AD biomarkers and neurocognitive status reached statistical significance in analyses for this subsample (Supplemental Material Table 3).

Exploratory analyses revealed a similar moderating pattern with mean cognitive performance as a dependent variable, with the exception of the non-significant moderation between t-Tau and neurocognitive status (Supplemental Material Table 4). Lastly, examining IICV in the context of the ATN model, the three-way interaction among Aβ, t-Tau, and p-Tau_181_ was significant in predicting both iSD and CoV (Supplemental Material Table 5). Post-hoc Tukey HSD tests revealed that the A+T+N+ group had a significantly lower demographically adjusted iSD z-score (i.e., higher IICV) than all other ATN groups (see Supplemental Material Figure 1 for visualization). In other words, those with A+T+N+ profiles demonstrated greater IICV than all other biomarker groups, even those with positive (+) status on one or two markers in the ATN model.

## Discussion

This study investigated the relation between AD biomarkers (CSF, PET) and IICV, as moderated by neurocognitive status. In line with our hypotheses, we found that individuals with cognitive impairment and elevated CSF and PET biomarkers of AD had elevated IICV, while no significant association emerged among cognitively intact individuals. Study results remained significant even after controlling for demographic covariates and were similar to the pattern observed from measures of central tendency. The current study validates the relationship of IICV with AD biomarker accumulation in cognitively impaired participants, as measured with CSF and amyloid PET with two separate samples from in the large NACC cohort. Consistent with prior research^1^, our findings suggest that IICV can be used to differentiate cognitively normal individuals from those with MCI and mild dementia. Further, our results show that increased IICV is related to both amyloid and tau and therefore may be a useful outcome measure for tracking clinical and biological disease progression or treatment response.

Empirical evidence thus far supports IICV as a harbinger of neurocognitive disorders, including both MCI and dementia^7^. IICV is sensitive to subtle neurocognitive changes due not only to AD but also other neurological conditions, including Parkinson’s disease^28^, Lewy Body disease^29^, frontotemporal dementia^30^, traumatic brain injury^31,32^, and human immunodeficiency virus^33,34^, but also to diseases and disorders that are not a direct insult on the central nervous system per se, such as breast cancer^35^. Although its sensitivity to multiple conditions suggests a low specificity in the context of differentiating clinical disorders^35^, the value of IICV rests in its sensitivity to brain pathology. Here, our current study linked IICV to AD biomarkers, including the accumulation of Aβ_1-42_, t-Tau, and p-Tau_181_ in CSF and in the brain as shown on PET scans. Such evidence is valuable in that most prior IICV studies focused on its correlation with clinical diagnoses while the current study directly validated IICV against an AD neurobiological process. Importantly, the findings were specific to those with neurocognitive impairment. Indeed, IICV was not meaningfully associated with AD biomarkers among cognitively healthy participants with our cross-sectional data analysis. However, based on previous findings, elevated IICV is likely a harbinger of neurocognitive disorders that can only be evaluated with longitudinal data^9^ although for a subset of those with elevated IICV, IICV may simply reflect normal ("benign") psychometric variability.

Regarding neural mechanisms of IICV, one popular perspective is that is that IICV likely reflects fronto-subcortical dysfunction and is a reflection of a failure of executive control^36,37^. In essence, consistent performance on a long battery of neuropsychological tests requires the ability to consistently marshal cognitive resources to maintain steady task performance over a prolonged period^4,38^. Neuroimaging studies concur with this point of view showing the role of neural correlates of executive functioning with IICV, including white matter integrity in superior longitudinal fasciculus^39^, body and genu of the corpus collosum, and anterior corona radiata as well as frontally mediated neural networks, including the default mode and the fronto-parietal executive control networks. Although typical AD is thought of as an amnestic presentation with mesial temporal involvement, alterations in frontally mediated neural networks have been described frequently in the literature^40^. Outcome measures in Alzheimer’s disease (AD) clinical trials most commonly include clinician-rated scales, like the CDR, or broad cognitive screeners, like the Mini Mental Status Exam. These measures often lack sensitivity to subtle cognitive impairments in the early stages of the disease^41^. To address these limitations, new measures have emerged, such as the preclinical Alzheimer’s cognitive composites (PACCs), which summarizes performance across a brief battery of neuropsychological tests. Though promising, such measures of global cognitive performance lack an underlying conceptual rationale for their relevance to AD. Thus, there continues to be a need for outcome measures that sensitively track AD progression and capitalize on our understanding of the underlying neuropathological changes of the disease, which underscores the importance of the current study. The current study found that among cognitively impaired individuals, as AD biomarkers increase, so too does IICV. That is, in the absence of AD biomarker assays, IICV may provide clinically meaningful information about disease progression that can complement traditional measures obtained from the neuropsychological assessment.

Importantly, results from the current study caution against the overgeneralization of our primary findings across ethno-racial groups. We found that among a subsample of Black and African American older adults, aside from p-Tau_181_, IICV was not related to any AD biomarkers, and the moderation by neurocognitive status was not statistically significant. Such findings should be interpreted in the context of our small sample size for Black and African American older adults with CSF data (n = 33) and those with neurocognitive impairment (n = 25). The null results may simply be due to insufficient statistical power. Nevertheless, our study does highlight the importance of future investigation of IICV in diverse cultural contexts^42^.

A particular strength of the current study is the use of demographically adjusted IICV scores. It is without question that neuropsychological tests are best interpreted with demographic adjustment^43^. However, this practice has not been routine in empirical studies on IICV or clinical evaluations despite its clear advantages. Prior studies showed that effects of IICV may be suppressed with raw IICV scores, specifically those generated based on parameters from the same sample and can only be uncovered with demographically adjusted IICV^34^. We argue the necessity to use demographically adjusted IICV instead of raw IICV for future studies for a better characterization of IICV that is unbiased against demographic disparity. Equally important, we also encouraged the use of IICV based on existing norms in clinical settings to better account for demographic factors and less reliant on qualitative clinical judgment^21^.

Limitations of the current study primarily centered around points of discussion in the IICV literature. First, whether the predictability of IICV differs by the neuropsychological tests that were used to calculate the index is unclear, thereby potentially limiting the generalizability of one single IICV study to IICVs calculated by different test batteries. Encouragingly, however, the relations between AD biomarkers and IICV were consistent across biomarkers, implying that the specific test battery used to calculate the index may be less of a concern than one would assume, though empirical evidence is required to draw such conclusion. Second, as examined in this study, there are two different methods of measuring IICV (i.e., iSD and CoV). Thus far, there is no consensus as to which one is superior to the other in terms of clinical utility^38^. In theory, CoV may be superior in that it mathematically controls the effect of the mean cognitive performance, which allows the examination of the effect of IICV above and beyond what can be expected by the measures of central tendency in isolation^21,23^. However, CoV comes at the cost of being less intuitive to interpret. Regardless, prior literature suggests that these two IICV indices are highly correlated and provide similar predictability in terms of neurological conditions. Our findings between the two IICV methods mostly concurred, with the sole exception that iSD appeared to better track AD biomarkers among the cognitively impaired individuals than CoV, which did not track with t-Tau and PET Aβ_1-42_. The decision on which index is more clinically meaningful remains a direction for future research. Third, the independent variable (AD biomarkers) and the moderators (neurocognitive status) in the regression analyses were not completely independent theoretically. However, all VIF values were within acceptable limits, indicating no statistical concern for multicollinearity. Future studies may benefit from mediational analyses to further examine the directionality of these associations—that is, whether elevations in AD biomarkers contribute to increased IICV, which in turn relates to neurocognitive diagnoses. Lastly, although the CSF Aβ42/40 ratio is recognized as a strong biomarker of AD pathology, we were unable to evaluate its association with IICV due to its absence in the NACC dataset. Future studies should explore this relation using datasets in which the Aβ42/40 ratio is available.

Overall, the current study not only replicated prior research showing that IICV differentiates cognitively impaired (incorporating both MCI and mild dementia) and cognitively healthy individuals but also discovered that increased IICV is linked to burden of amyloid-beta, tau and neurodegeneration in both CSF and with amyloid PET. The amyloid PET and the CSF data were independent cohorts within the NACC database and therefore collectively reached the same conclusion, replicating our findings. These results suggest that IICV may be a useful diagnostic tool complementing existing neuropsychological measures of central tendency. Additionally, IICV may be an appropriate AD disease outcome measure in both observational studies and clinical trials, specifically for disease modifying therapies among cognitively impaired individuals. Given small sample size and possible inadequate power, findings were not replicated in a subsample of Black and African Americans which highlights the need for further investigation on the relation between AD biomarkers and IICV among ethno-racially diverse populations. Lastly, the current study suggests the importance of using demographically adjusted IICV in research settings considering its clinical advantages.

## Data Availability

All data are available online through the National Alzheimers Coordinating Center (https://www.naccdata.org/)

